# Test sensitivity is secondary to frequency and turnaround time for COVID-19 surveillance

**DOI:** 10.1101/2020.06.22.20136309

**Authors:** Daniel B. Larremore, Bryan Wilder, Evan Lester, Soraya Shehata, James M. Burke, James A. Hay, Milind Tambe, Michael J. Mina, Roy Parker

**Author notes:** These authors contributed equally (Michael J. Mina, Roy Parker).

## Abstract

The COVID-19 pandemic has created a public health crisis. Because SARS-CoV-2 can spread from individuals with pre-symptomatic, symptomatic, and asymptomatic infections, the re-opening of societies and the control of virus spread will be facilitated by robust surveillance, for which virus testing will often be central. After infection, individuals undergo a period of incubation during which viral titers are usually too low to detect, followed by an exponential viral growth, leading to a peak viral load and infectiousness, and ending with declining viral levels and clearance. Given the pattern of viral load kinetics, we model surveillance effectiveness considering test sensitivities, frequency, and sample-to-answer reporting time. These results demonstrate that effective surveillance depends largely on frequency of testing and the speed of reporting, and is only marginally improved by high test sensitivity. We therefore conclude that surveillance should prioritize accessibility, frequency, and sample-to-answer time; analytical limits of detection should be secondary.

## Introduction

Successful surveillance testing of SARS-CoV-2 depends on understanding both the dynamics of spread between individuals and the dynamics of the virus within the human body. Critically, the ability of SARS-CoV-2 to spread from individuals who are pre-symptomatic, symptomatic, or essentially asymptomatic [1,2, 3] means that diagnosis and isolation based on symptoms alone will be unable to prevent ongoing spread [4, 5]. As a consequence, the use of surveillance testing to identify infectious individuals presents one possible means to break enough transmission chains to suppress the ongoing pandemic and reopen societies, with or without a vaccine.

The reliance on testing as a means to safely reopen societies has placed a microscope on the analytical sensitivity of virus assays, with a gold-standard of quantitative real-time polymerase chain reaction (qPCR). These assays have analytical limits of detection that are usually within around 10^3^ viral RNA copies per ml (cp/ml) [6]. However, qPCR remains expensive and as a laboratory based assay often have sample-to-result times of 24-48 hours. New developments in SARS-CoV-2 diagnostics have the potential to reduce cost significantly, allowing for expanded testing or greater frequency of testing and can reduce turnaround time to minutes [7, 8, 9]. These assays however largely do not meet the gold standard for analytical sensitivity, which has encumbered the widespread use of these assays [10].

Three features of the viral increase, infectivity, and decline during SARS-CoV-2 infection led us to hypothesize that there might be minimal differences in effective surveillance using viral detection tests of different sensitivities, such as RT-qPCR with a limit of detection (LOD) at 10^3^ cp/ml [6] compared to often cheaper or faster assays with higher limits of detection (i.e., around 10^5^ cp/ml [7, 8, 9]) such as point-of-care nucleic acid LAMP and rapid antigen tests (Figure 1A). First, since filtered samples collected from patients displaying less than 10^6^ N or E RNA cp/ml contain minimal or no measurable infectious virus [11, 12, 13], either class of test should detect individuals who are currently infectious. The absence of infectious particles at viral RNA concentrations < 10^6^ cp/ml is likely due to (i) the fact that the N and E RNAs are also present in abundant subgenomic mRNAs, leading to overestimation of the number of actual viral genomes by ~100-1000X [14], (ii) technical artifacts of RT-PCR at Ct values > 35 due to limited template [15, 16], and (iii) the production of non-infectious viral particles as is commonly seen with a variety of RNA viruses [17]. Second, during the exponential growth of the virus, the time difference between 10^3^ and 10^5^ cp/ml is short, allowing only a limited window in which only the more sensitive test could diagnose individuals. For qPCR, this corresponds to the time required during viral growth to go from Ct values of 40 to ~34. While this time window for SARS-CoV-2 is not yet rigorously defined [18], for other respiratory viruses such as influenza, and in ferret models of SARS-CoV-2 transmission, it is on the order of a day [19, 20]. Finally, high-sensitivity screening tests, when applied during the viral decline accompanying recovery, are unlikely to substantially impact transmission because such individuals detected have low, if any, infectiousness [14]. Indeed, a recent review by Cevik et al [18] notes that no study to date has successfully cultured live virus more than 9 days after the onset of symptoms.

**Figure 1:**
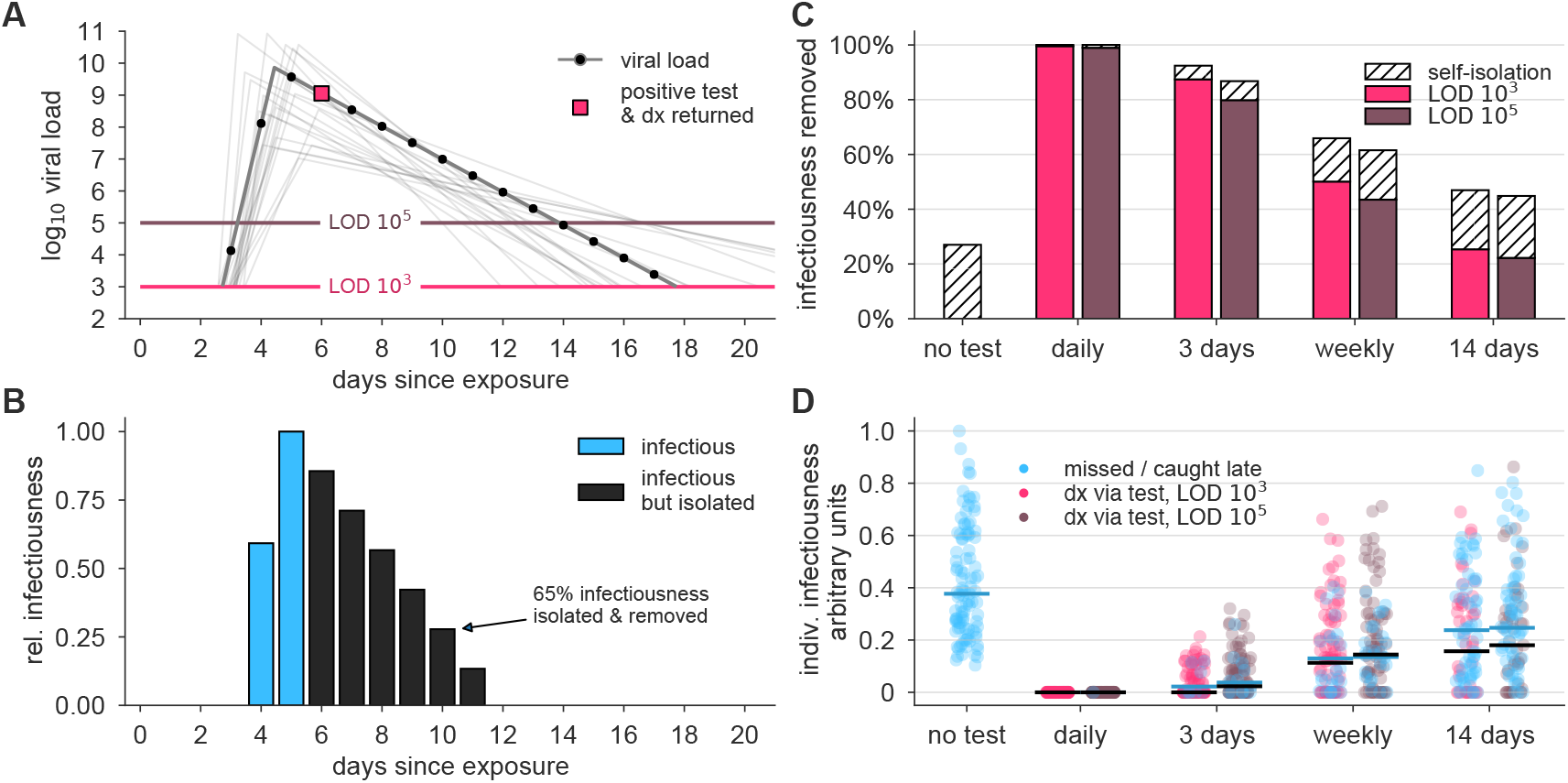
Surveillance testing effectiveness depends on frequency. (A) An example viral load trajectory is shown with LOD thresholds of two tests, and a hypothetical positive test on day 6, two days after peak viral load. 20 other stochastically generated viral loads are shown to highlight trajectory diversity (light grey; see Methods). (B) Relative infectiousness for the viral load shown in panel A pre-test, totaling 35% (blue) and post-isolation, totaling 65% (black). (C) Surveillance programs using tests at LODs of 10^3^ and 10^5^ at frequencies indicated were applied to 10,000 individuals’ trajectories of whom 35% would undergo symptomatic isolation near their peak viral load if they had not been tested and isolated first. Total infectiousness removed during surveillance (colors) and self isolation (hatch) are shown for surveillance as indicated, relative to total infectiousness with no surveillance or self-isolation. (D) The impact of surveillance on the infectiousness of 100 individuals is shown for each surveillance program and no testing, as indicated, with each individual colored by test if their infection was detected during infectiousness (medians, black lines) or colored blue if their infection was missed by surveillance or detected positive *after* their infectious period (medians, blue lines). Units are arbitrary and scaled to the maximum infectiousness of sampled individuals.

## Results

### Impact of surveillance on individuals

To examine how surveillance testing would reduce the average infectiousness of individuals, we first modeled the viral loads and infectiousness curves of 10,000 simulated individuals using the predicted viral trajectories of SARS-CoV-2 infections based on key features of latency, growth, peak, and decline identified in the literature (Figure 1A; see Methods). Accounting for these within-host viral kinetics, we calculated what percentage of their total infectiousness would be removed by surveillance and isolation (Figure 1B) with tests at LOD of 10^3^ and 10^5^, and at different frequencies. Here, infectiousness was taken to be proportional to the logarithm of viral load in excess of 10^6^ cp/ml (with alternative assumptions addressed in sensitivity analyses; see Supplemental Materials), consistent with the observation that pre-symptomatic patients are most infectious just prior to the onset of symptoms [21], and evidence that the efficiency of viral transmission coincides with peak viral loads, which was also identified during the related 2003 SARS outbreak [22, 23]. We considered that 35% of patients would undergo symptomatic isolation within three days of their peak viral load if they had not been tested and isolated first, and 65% would have sufficiently mild or no symptoms such that they would not isolate unless they were detected by surveillance testing. Based on recent results, we modeled asymptomatic and symptomatic infections as having the same initial viral loads [1, 24, 25, 26], but with faster clearance among asymptomatics [24, 26, 27, 28, 29] (see Methods). This analysis demonstrated that there was little difference in averting infectiousness between the two classes of test. Dramatic reductions in total infectiousness of the individuals were observed by testing daily or every third day, ~ 65% reduction when testing weekly, and < 50% under biweekly testing (Figure 1C). Because viral loads and infectiousness vary across individuals, we also analyzed the impact of different surveillance regimens on the distribution of individuals’ infectiousness, revealing that more sporadic testing leads to an increased likelihood that individuals will test positive after they are no longer infectious or be missed by testing entirely (Figure 1D).

### Impact of surveillance on a population

Above, we assumed that each infection was independent. To investigate the effects of surveillance testing strategies at the population level, we used simulations to monitor whether epidemics were contained or became uncontrolled, while varying the frequencies at which the test was administered, ranging from daily testing to testing every 14 days, and considering tests with LOD of 10^3^ and 10^5^, analogous to RT-qPCR and RT-LAMP / rapid antigen tests, respectively. We integrated individual viral load trajectories into two different epidemiological models to ensure that important observations were independent of the specific modeling approach. The first model is a simple fully mixed model representing a population of 20,000, similar to a large university setting, with a constant rate of external infection approximately equal to one new import per day. The second model is a previously described agent-based model with both within-household and age-stratified contact structure based on census microdata in a city representative of New York City [30], which we initialized with 100 cases without additional external infections. Individual viral loads were simulated for each infection, and individuals who received a positive test result were isolated, but contact tracing and monitoring was not included to more conservatively estimate the impacts of surveillance alone [31, 32]. Model details and parameters are fully described in Methods.

We observed that a surveillance program administering either test with high frequency limited viral spread, measured by both a reduction in the reproductive number *R* (Figures 2A and B; see Methods for calculation procedure) and by the total infections that persisted in spite of different surveillance programs, expressed relative to no surveillance (Figures 2C and D). Testing frequency was found to be the primary driver of population-level epidemic control, with only a small margin of improvement provided by using a more sensitive test. Direct examination of simulations showed that with no surveillance or biweekly testing, infections were uncontrolled, whereas surveillance testing weekly with either LOD = 10^3^ or 10^5^ effectively attenuated surges of infections (examples shown in Figure S1).

**Figure 2:**
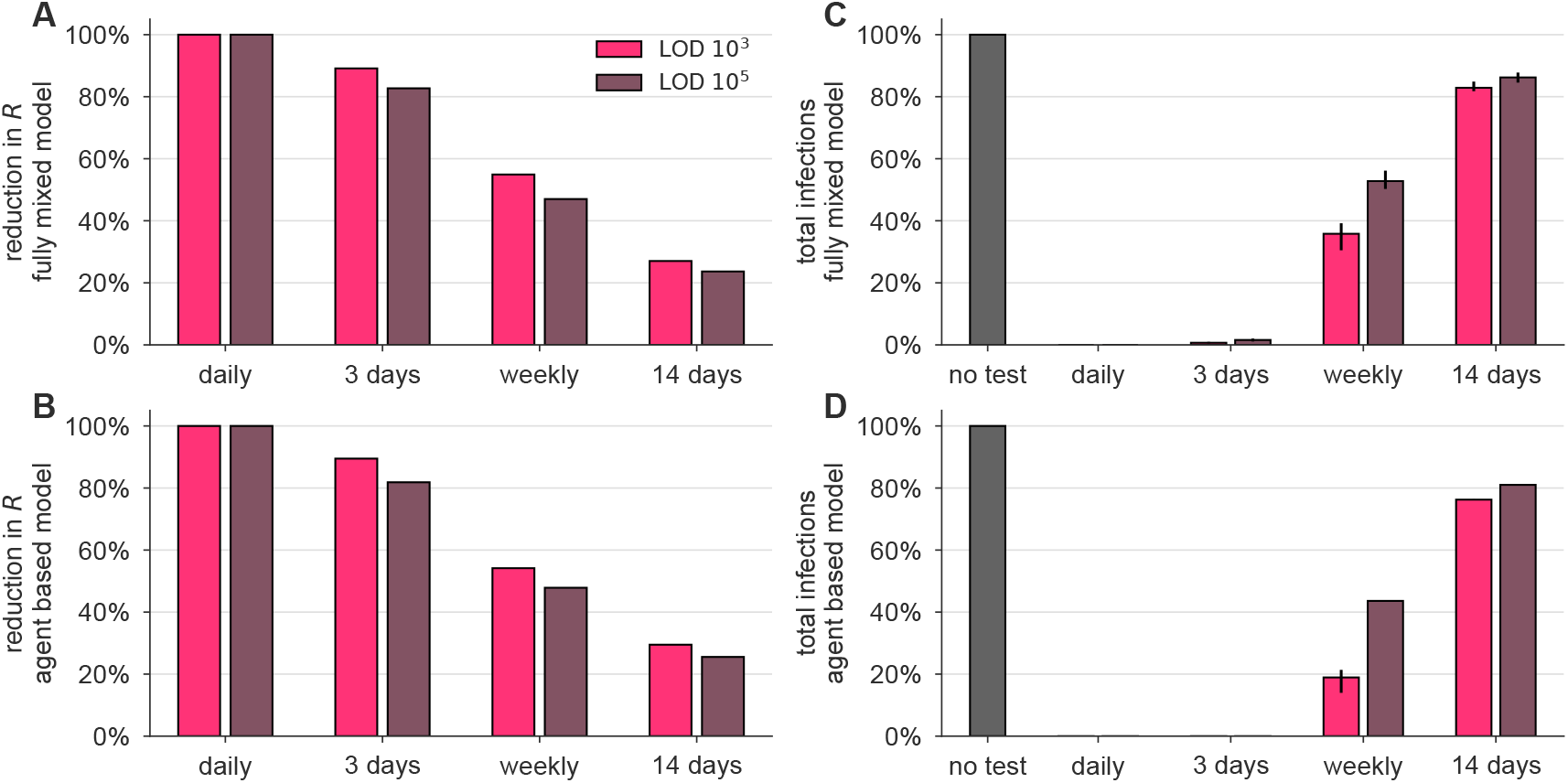
Surveillance testing affects disease dynamics. Both the fully-mixed compartmental model (top row) and agent based model (bottom row) are affected by surveillance programs. (A, B) More frequent testing reduces the effective reproductive number *R*, shown as the percentage by which *R*_0_ is reduced, 100 x (*R*_0_ - *R)/R*_0_. Values of *R* were estimated from 50 independent simulations of dynamics (see Methods). (C, D) Relative to no testing (grey bars), surveillance suppresses the total number of infections in both models when testing every day or every three days, but only partially mitigates total cases for weekly or bi-weekly testing. Error bars indicate inner 95% quantiles of 50 independent simulations each.

The relationship between test sensitivity and the frequency of testing required to control outbreaks in both the fully mixed model and the agent-based model generalize beyond the examples shown in Figure 2 and are also seen at other testing frequencies, sensitivities, and asymptomatic fractions. We simulated both models at LODs of 10^3^, 10^5^, and 10^6^, and for testing ranging from daily to every 14 days. For those, we measured each surveillance policy’s impact on total infections (Figure S2A and B) and on R (Figure S2C and D). In Figure 2, we modeled infectiousness as proportional to log_10_ of viral load. To address whether these finding are sensitive to this modeled relationship, we performed similar simulations with infectiousness proportional to viral load (Figure S3), or uniform above 10^6^/ml (Figure S4). We found that results were robust to these large variations in the modeled relationship between infectiousness and viral load. To further address whether our results depended on the exact 35% fraction of individuals assumed to be behaviorally symptomatic, we performed sensitivity analyses with fewer (20%) or more (50%) symptomatic individuals and found no meaningful difference in results (Figure S5).

### Impact of delayed test results

An important variable in surveillance testing is the time between a test’s sample collection and the reporting of a diagnosis. To examine how time to reporting affected epidemic control, we re-analyzed both the reduction in individuals’ infectiousness, as well as the epidemiological simulations, comparing the results of instantaneous reporting (reflecting a rapid point-of-care assay), one day delay, and two day delay (Figure 3A and B). Delays in reporting dramatically decreased the reduction in infectiousness in individuals as seen by the total infectiousness removed (Figure 3C), the distribution of infectiousness in individuals (Figure 3D), or the dynamics of the epidemiological models (Figure 4). This result was robust to the modeled relationship between infectiousness and viral load in both simulation models and for various test sensitivities and frequencies (Figure S6). These results highlight that delays in reporting lead to dramatically less effective control of viral spread and emphasize that fast reporting of results is critical in any surveillance testing. These results also reinforce the relatively smaller benefits of improved limits of detection.

**Figure 3:**
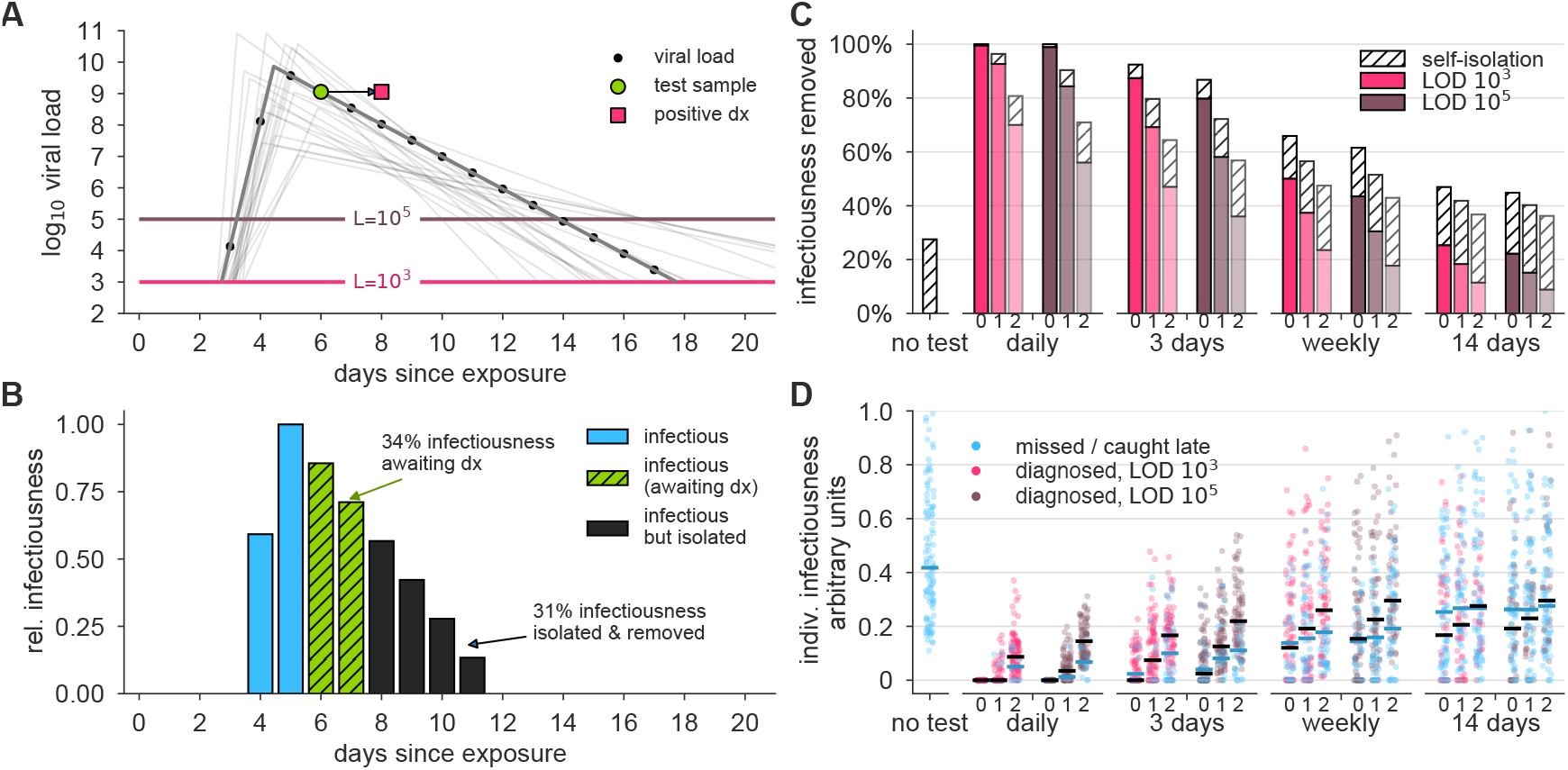
Effectiveness of surveillance testing is compromised by delays in reporting. (A) An example viral load trajectory is shown with LOD thresholds of two tests, and a hypothetical positive test on day 6, but with results reported on day 8. 20 other stochastically generated viral loads are shown to highlight trajectory diversity (light grey; see Methods). (B) Relative infectiousness for the viral load shown in panel A pre-test (totaling 35%; blue) and posttest but pre-diagnosis (totaling 34%; green), and post-isolation (totaling 31%; black). (C) Surveillance programs using tests at LODs of 10^3^ and 10^5^ at frequencies indicated, and with results returned after 0, 1, or 2 days (indicated by small text beneath bars) were applied to 10,000 individuals trajectories of whom 35% were symptomatic and self-isolated after peak viral load if they had not been tested and isolated first. Total infectiousness removed during surveillance (colors) and self isolation (hatch) are shown, relative to total infectiousness with no surveillance or self-isolation. Delays substantially impact the fraction of infectiousness removed. (D) The impact of surveillance with delays in returning diagnosis of 0, 1, or 2 days (small text beneath axis) on the infectiousness of 100 individuals is shown for each surveillance program and no testing, as indicated, with each individual colored by test if their infection was detected during infectiousness (medians, black lines) or colored blue if their infection was missed by surveillance or diagnosed positive *after* their infectious period (medians, blue lines). Units are arbitrary and scaled to the maximum infectiousness of sampled individuals.

**Figure 4:**
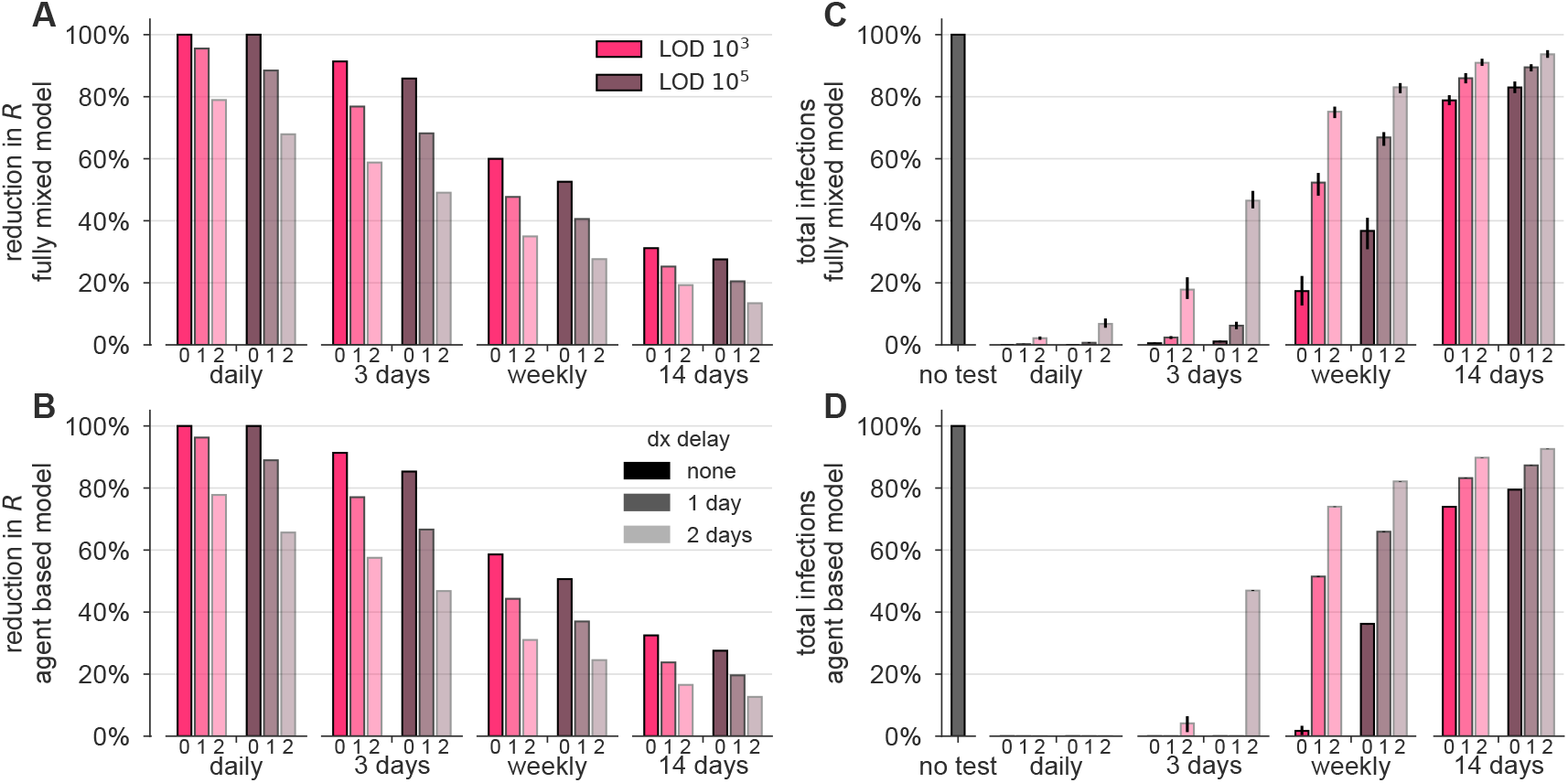
Delays in reporting decrease the epidemiological impact of surveillance-driven isolation. The effectiveness of surveillance programs are dramatically diminished by delays in reporting in both the fully-mixed compartmental model (top row) and agent based model (bottom row). (A, B) The impact of surveillance every day, 3 days, weekly, or biweekly, on the reproductive number R, calculated as 100 x (*R*_0_ - *R)/R*_0_, is shown for LODs 10^3^ and 10^5^ and delays of 0, 1, or 2 days (small text below axis). Values of R were estimated from 50 independent simulations of dynamics (see Methods). (C, D) Relative to no testing (grey bars), surveillance suppresses the total number of infections in both models when testing every day or every three days, but delayed results lead to only partial mitigation of total cases, even for testing every day or 3 days. Error bars indicate inner 95% quantiles of 50 independent simulations each.

### Generality of findings to changes in modeling assumptions

Communities vary in their transmission dynamics, due to difference in rates of imported infections and in the basic reproductive number *R*_0_, both of which will influence the frequency and sensitivity with which surveillance testing must occur. We performed two analyses to illustrate this point. First, we varied the rate of external infection in our fully mixed model, and confirmed that when the external rate of infection is higher, more frequent surveillance is required to prevent outbreaks (Figure S7A). Second, we varied the reproductive number *R*_0_ between infected individuals in both models, and confirmed that at higher *R*_0_, more frequent surveillance is also required (Figure S7B and C). This may be relevant to institutions like college campuses or military bases wherein frequent classroom setting or dormitory living are likely to increase contact rates. Thus, the specific strategy for successful surveillance will depend on the current community infection prevalence and transmission rate.

The generality of our findings to different epidemiological parameters (Figure S7), relationships between viral load and infectiousness (Figures S3 and S4), and proportion of symptomatic individuals (Figure S5) led us to ask whether a more general mathematical formula could predict R without requiring epidemiological simulation. We derived such a formula (Supplemental Text S1) and found that its predicted values of R were nearly perfectly correlated with simulation-estimated values (Pearson’s r = 0.998, *p <* 10^-6^; Figure S8), providing a mathematical alternative to simulation-based sensitivity analyses.

### Surveillance testing to mitigate an ongoing epidemic

The impact of surveillance testing on transmission dynamics led us to hypothesize that surveillance testing could be used as an active tool to mitigate an ongoing epidemic. To test this idea, we simulated an outbreak situation using both the fully-mixed and agent-based models but with three additional conditions. First, we assumed that in an ongoing pandemic, other mitigating interventions would cause the reproductive number to be lower, though nevertheless larger than one. Second, we considered that not all individuals would want to or be able to participate in a SARS-CoV-2 surveillance program. Third, we assumed that the collection of samples for testing, if performed on a large scale, could result in imperfect sample collection, causing an increase in the false negative rate, independent of an assay’s analytical sensitivity. These modifications are fully described in Methods.

We simulated epidemics in which surveillance testing began only at the point when uncontrolled infections reached 4% prevalence. Based on results from our previous analyses, we considered a less sensitive but rapid test with LOD 10^5^ cp/ml and a zero-day delay in results, and further assumed that 10% of would-be positive samples would be negative due to improper sample collection. We then examined scenarios of testing every 3 days and every 7 days, with either 50% or 75% of individuals participating, starting from a partially mitigated *R*_0_ = 1.5. We found that surveillance testing of 75% of individuals every 3 days was sufficient to drive the epidemic toward extinction within 6 weeks and reduce cumulative incidence by 88%, and that other combinations also had successful but less rapid mitigating impacts, particularly when compared with no intervention (Figure 5). Notably, even weekly testing with 50% participation was able to reduce the peak and length of the outbreak, illustrating how even partial surveillance testing using a test with 100X lower molecular sensitivity than PCR can have public health benefits when used frequently (Figure 5). Repeating these simulations using a test with LOD 10^6^ led to similar results (Figure S9). To further generalize these results, we modified our mathematical formula to predict the impacts of per-individual test refusal and per-test sampling-related sensitivity on the reproductive number R (see Supplemental Text S1).

**Figure 5:**
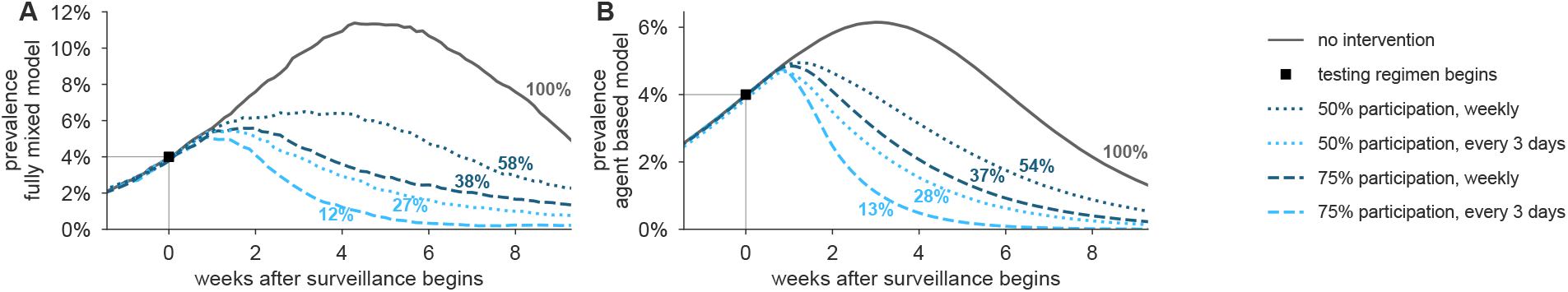
Surveillance testing suppresses an ongoing epidemic. Widespread testing and isolation of infected individuals drives prevalence downward for both (A) the fully-mixed compartmental model and (B) the agent based model. Time-series of prevalence, measured as the total number of infectious individuals, are shown for no intervention (solid) and surveillance testing scenarios (various dashed; see legend). Surveillance testing began only when prevalence reached 4% (box), and time series are shifted such that testing begins at t = 0. Scenarios show the impact of a test with LOD 10^5^, no delay in results, and with 10% of samples assumed to be incorrectly collected (and therefore negative) to reflect decreased sensitivity incurred at sample collection in a mass testing scenario. Annotations show total number of post-intervention infections, as a percentage of the no-intervention scenario, labeled as 100%. See Fig. S9 for identical simulations using a test with LOD 10^6^.

## Discussion

Our results lead us to conclude that surveillance testing of asymptomatic individuals can be used to limit the spread of SARS-CoV-2. However, our findings are subject to a number of limitations. First, the sensitivity of a test may depend on factors beyond LOD, including manufacturer variation and improper clinical sampling [33], though the latter may be ameliorated by different approaches to sample collection, such as saliva-based testing [34]. Second, the exact performance differences between testing schemes will depend on whether our model truly captures viral kinetics and infectiousness profiles [21], particularly during the acceleration phase between exposure and peak viral load. Continued clarification of these within-host dynamics would increase the impact and value of this, and other [31, 32, 35, 36] modeling studies. Finally, we modeled participation in surveillance testing regimens (or refusal thereof) as statistically independent between individuals, but health-related behaviors have been shown to be socially [37] and geographically [38, 39] correlated. Clustered refusal of surveillance testing, or refusal to isolate upon testing positive, could present challenging barriers to implementation.

Our findings show that the impact of surveillance testing can be expressed as a reduction of the reproductive number R. By mapping a given testing regimen to a reduction in R, the impact of testing regimen can be approximated and generalized without complicated simulations. For instance, one could estimate the maximum allowable turnaround time delays, or the minimum testing frequency required to bring *R* below one, based on user-specified and scenario-specific assumptions. To facilitate such generalizations and scenario planning, open-source calculation tools accompany this manuscript.

A critical point is that the requirements for surveillance testing are distinct from clinical testing. Clinical diagnoses target symptomatic individuals, need high accuracy and sensitivity, and are not limited by cost. Because they focus on symptomatic individuals, those individuals can isolate such that a diagnosis delay does not lead to additional infections. In contrast, results from the surveillance testing of asymptomatic individuals need to be returned quickly, since even a single day diagnosis delay compromises the surveillance program’s effectiveness. Indeed, at least for viruses with infection kinetics similar to SARS-CoV-2, we find that speed of reporting is much more important than sensitivity, although more sensitive tests are nevertheless somewhat more effective.

The difference between clinical and surveillance testing highlights the need for additional tests to be approved and utilized for surveillance. Such tests should not be held to the same degree of sensitivity as clinical tests, in particular if doing so encumbers rapid deployment of faster cheaper SARS-CoV-2 assays. We suggest that the FDA, other agencies, or state governments, encourage the development and use of alternative faster and lower cost tests for surveillance purposes, even if they have poorer limits of detection. If the availability of point-of-care or self-administered surveillance tests leads to faster turnaround time or more frequent testing, our results suggest that they would have high epidemiological value.

Our modeling suggests that some types of surveillance will subject some individuals to unnecessary quarantine days. For instance, the infrequent use of a sensitive test will not only identify (i) those with a low viral load in the beginning of the infection, who must be isolated to limit viral spread, but (ii) those in the recovery period, who still have detectable virus or RNA but are below the infectious threshold [13, 14]. Isolating this second group of patients will have no impact on viral spread but will incur costs of isolation, as would the isolation of individuals who received a false positive test result due to imperfect test specificity. The use of serology, repeat testing 24 or 48 hours apart, or some other test, to distinguish low viral load patients on the upslope of infection from those in the recovery phase could allow for more effective quarantine decisions.

## Materials and Methods

### Viral Loads

Viral loads were drawn from a simple viral kinetics model intended to capture (1) a variable latent period, (2) a rapid growth phase from the lower limit of PCR detectability to a peak viral load, (3) a slower decay phase, and (4) prolonged clearance for symptomatic infections vs asymptomatic infections. These dynamics were based on the following observations.

Latent periods prior to symptoms have been estimated to be around 5 days [40]. Latent periods prior to detection via virological tests at secondary sites of replication or shedding have been estimated to be up to 4 days [41], corresponding to a latent or eclipse phase observed with other viruses [42]. Viral load appears to peak prior to symptom onset [21], and peaks within 2 days of challenge in a macaque model [43, 44], though it should be noted that macaque challenge doses were high. Viral load decreases monotonically from the time of symptom onset [21, 45, 46, 47, 48], but may be high and detectable 3 or more days before symptom onset [1, 49]. Peak viral loads are difficult to measure due to lack of prospective sampling studies of individuals prior to exposure and infection, but viral loads have been reported in the range of 0 (10^4^) to 0(10^9^) copies per ml [12, 47, 48]. Viral loads appear to become undetectable by PCR within 3 weeks of symptom onset [45, 48, 50], but detectability and timing may differ depending on the degree or presence of symptoms [50, 51]. The majority of studies reviewed by Cevik et al [18] found initial viral loads to be similar between symptomatic and asymptomatic infections [1, 24, 25, 26], but viral clearance was significantly and substantially faster among asymptomatic infections [24, 26, 27, 28, 29]. Finally, we note that the general understanding of viral kinetics may vary depending on the mode of sampling, as demonstrated via a comparison between sputum and swab samples [12]. For a comprehensive review of viral load dynamics, duration of shedding, and infectiousness, see Ref. [18].

To mimic growth and decay, log_10_ viral loads were specified by a continuous piecewise linear “hinge” function, specified uniquely with three control points: *(t_0_*, 3), (*t*_peak_, *V*_peak_),(*t_f_*, 6) (Figure 6A; green squares). The first point represents the time at which an individual’s viral load first crosses 10^3^, with *t*_0_ ~ unif[2.5, 3.5], measured in days since exposure. The second point represents the peak viral load. Peak height was drawn *V*_peak_ ~ unif[7,11], and peak timing was drawn with respect to the start of the exponential growth phase, *t*_peak_ − *t*_0_ ~ 0.5 + gamma(1.5) with a maximum of 3. The third point represents the time at which an individual’s viral load crosses beneath the 10^6^ threshold, at which point viral loads no longer cause active cultures in laboratory experiments [11, 12, 13, 18]. For asymptomatic infections, this point was drawn with respect to peak timing, *t_f_* − *t*_peak_ ~ unif[4, 9]. For symptomatic infections, a symptom onset time was first drawn with respect to peak timing, *t*_symptoms_ − *t*_peak_ ~ unif[0, 3], and then the third control point was drawn with respect to symptom onset, *t_f_* − *t*_symptoms_ ~ unif[4, 9]. Thus, symptomatic trajectories are systematically longer, in both duration of infectiousness (see below) and duration of viral shedding, reflecting the documented prolonged clearance and relationship with viral culture experiments (Figure 6B; red circles). In simulations, each viral load’s parameters were drawn independently of others, and the continuous function described here was evaluated at 28 integer time points (Figure 6; black dots) representing a four week span of viral load values.

**Figure 6:**
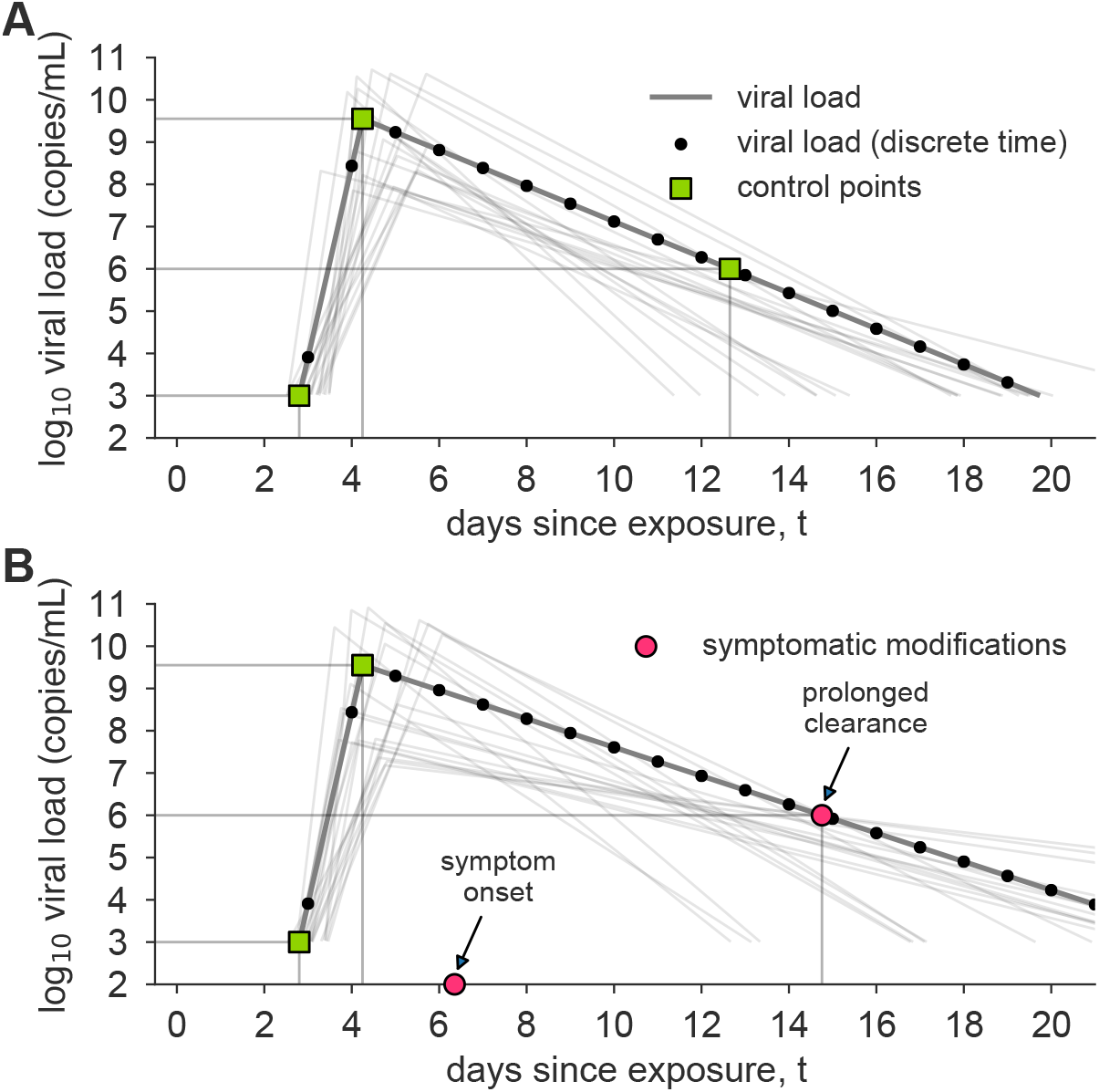
Example asymptomatic and symptomatic viral loads with model control points. Examples of model viral loads (lines) and corresponding stochastically drawn control points (squares, circles) are shown for (A) an asymptomatic viral load trajectory and (B) a symptomatic viral load trajectory. Because simulations took place in discrete time, black dots show points at which this example viral load would have been sampled. Light grey lines show 20 alternative trajectories in each panel to illustrate the diversity of viral loads drawn from the simple model. Red circles indicate the control points which are modified in symptomatic trajectories to account for symptom onset and prolonged time till clearance.

### Infectiousness

Infectiousness F was assumed to be directly related to viral load V in one of three ways. In the main text, each individual’s relative infectiousness was proportional log_10_ of viral load’s excess beyond 10^6^, i.e. *F* ∝ log_10_(*V*) − 6. In the supplementary sensitivity analyses, we investigated two opposing extremes. To capture a more extreme relationship between infectiousness and viral load, we considered *F* to be directly proportional to viral load’s excess above 10^6^, i.e. 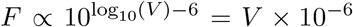, and to capture a more extreme relationship, but in the opposing direction, we considered *F* to simply be a constant when viral load exceeded 10^6^, i.e. 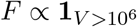. We call these three functions *log-proportional, proportional*, and *threshold* throughout the text and supplemental materials.

We note that a comprehensive review of viral loads, shedding, and infectiousness [18] found that across the surveyed literature, no virus was able to be cultured beyond 9 days post-symptoms. Thus, the choice of the final control point in our symptomatic viral load model (Figure 6B), which corresponds to the latest time at which an individual is infectious, is at most 9 days post-symptom onset.

Recently, He et al [21] published an analysis of infectiousness relative to symptom onset which was corrected by Bonhoeffer et al (see [21] for details). Among our infectiousness functions, this inferred relationship bears the greatest similarity, over time, to the log-proportional infectiousness function, as visualized in Figs. 1 and 3. The proportional and threshold models therefore represent one of many types of sensitivity analysis. Results for those models can be found in Figures S3, S4, and S6.

In all simulations, the value of the proportionality constant implied by the infectiousness functions above was chosen to achieve the targeted value of *R*_0_ for that simulation, and confirmed via simulation as described below.

### Disease Transmission Models

#### Overview

Two models were used to simulate SARS-CoV-2 dynamics, both based on a typical compartmental framework. The first model was a fully-mixed model of *N* = 20, 000 individuals with all-to-all contact structure, zero initial infections, and a constant 1/*N* per-person probability of becoming infected from an external source. This model could represent, for instance, a large college campus with high mixing, situated within a larger community with low-level disease prevalence. The second model was an agent-based model of *N* = 8.4 million agents representing the population and contact structure of New York City, as previously described [30]. Contact patterns were based on a combination of individual-level household contacts drawn from census microdata and age-stratified contact matrices which describe outside of household contacts. This model was initialized with 100 initial infections and no external sources of infection.

Both the fully-mixed and agent-based models tracked discrete individuals who were Susceptible (*S*), Infected (*I*), Recovered (*R*), Isolated (*Q*), and Self-Isolated (*SQ*) at each discrete one day timestep. Upon becoming infected (*S* → *I*), a viral load trajectory *V(t*) was drawn which included a latent period, growth, and decay. Each day, an individual’s viral load trajectory was used to determine whether their diagnostic test would be positive if administered, as well as their infectiousness to susceptible individuals. Based on a schedule of testing each person every *D* days, if an individual happened to be tested on a day when their viral load exceeded the limit of detection *L* of the test, their positive result would cause them to isolate (*I* → *Q*), but with the possibility of a delay in turnaround time. A fraction 1 − *f* of individuals self-isolate on the day of symptom onset, which occurs 0 to 3 days after peak viral load, to mimic symptom-driven isolation (*I* → *SQ*), with *f* = 0.65 for both models, with *f* = 0.8 and *f* = 0.5 explored in sensitivity analyses (Figure S5). Presymptomatic individuals were isolated prior to symptom onset only if they received positive test results. When an individual’s viral load dropped below 10^3^, that individual recovered (*I*, *Q, SQ → R*). Details follow.

#### Testing, Isolation, and Sample-to-Answer Turnaround Times

All individuals were tested every D days, so that they could be moved into isolation if their viral load exceeded the test’s limit of detection *V(t) > L*. Each person was deterministically tested exactly every D days, but testing days were drawn uniformly at random such that not all individuals were tested on the same day. To account for delays in returning test results, we included a sample-to-answer turnaround time *T*, meaning that an individual with a positive test on day t would isolate on day *t + T*.

#### Transmission, Population Structure, and Mixing Patterns: Fully-mixed model

Simulations were initialized with all individuals susceptible, *S = N*. Each individual was chosen to be symptomatic independently with probability *f*, and each individual’s first test day (e.g. the day of the week that their weekly test would occur) was chosen uniformly at random between 1 and *D*. Relative infectiousness was scaled up or down to achieve the specified *R*_0_ in the absence of any testing policy, but inclusive of any assumed self-isolation of symptomatics.

In each timestep, those individuals who were marked for testing that day were tested, and a counter was initialized to *T*, specifying the number of days until that individual received their results. Next, individuals whose test results counters were zero were isolated, *I* → *Q*. Then, symptomatic individuals whose viral load had declined relative to the previous day were self-isolated, *I* → *SQ*. Next, each susceptible individual was spontaneously (externally) infected independently with probability 1/*N*, *S* → *I*. Then, all infected individuals contacted all susceptible individuals, with the probability of transmission based on that day’s viral load *V (t*) for each person and the particular infectiousness function, described above, *S* → *I*.

To conclude each time step, individuals’ viral loads and test results counters were advanced, with those whose infectious period had completely passed moved to recovery, *I*, *Q*, *SQ* → *R*.

#### Transmission, Population Structure, and Mixing Patterns: Agent-based model

The agent-based model added viral kinetics and testing policies (as described above) to an existing model for SARS-CoV-2 transmission in New York City. A full description of the agent-based model is available [30]; here we provide an overview of the relevant transmission dynamics.

Simulations were initialized with all individuals susceptible, except for 100 initially infected individuals, *S* = *N* − 100. As in the fully-mixed model, each individual’s test day was chosen uniformly at random and relative infectiousness was scaled to achieve the specified *R*_0_.

In each timestep, those individuals who were marked for testing that day were tested, and a counter was initialized to T, specifying the number of days until that individual received their results. Next, individuals whose test results counters were zero were isolated, *I* → *Q*. There was no self-isolation in this model (and accordingly, the model did not label individuals as symptomatic or asymptomatic).

Then, transmission from infected individuals to susceptible individuals was simulated both within and outside households. To model within-household transmission, each individual had a set of other individuals comprising their household. Household structures, along with the age of each individual, were sampled from census microdata for New York City [52]. The probability for an infectious individual to infect each of their household members each day was determined by scaling the relative infectiousness values to match the estimated secondary attack rate for close household contacts previously reported in case cluster studies [53].

Outside of household transmission was simulated using age-stratified contact matrices, which describe the expected number of daily contacts between an individual in a given age group and those in each other age group. Each infectious individual of age i drew Poisson(*M_ij_*) contacts with individuals in age group *j*, where *M* is the contact matrix. The contacted individuals were sampled uniformly at random from age group *j*. We use a contact matrix for the United States estimated by [54]. Each contact resulted in infection, *S → I*, with probability proportional to the relative infectiousness of the infected individual on that day, scaled to obtain the specified value of *R*_0_.

To conclude each time step, individuals’ viral loads and test results counters were advanced, with those whose infectious period had completely passed moved to recovery, *I,Q → R*.

#### Calibration to achieve targeted *R*_0_ and estimation of *R*

As a consistency check, each simulation’s *R*_0_ was estimated as follows, to ensure that simulations were properly calibrated to their intended values. Note that to vary *R*_0_, the proportionality constant in the function that maps viral load to infectiousness need only be adjusted up or down. In a typical SEIR model, this would correspond to changing the infectiousness parameter which governs the rate at which I-to-S contacts cause new infections *β*.

For the fully-mixed, the value of *R*_0_ was numerically estimated by running single-generation simulations in which a 50 infected individual were placed in a population of *N −* 50 others. The number of secondary infections from those initially infected was recorded and used to directly estimate *R*_0_.

For the agent-based model, the value of *R*_0_ depends on the distribution of infected agents due to stratification by age and household. We numerically estimate *R*_0_ by averaging over the number of secondary infections caused by each agent who was infected in the first 15 days of the simulation (at which point the population is still more than 99.99% susceptible).

Estimations of R proceeded exactly as estimations of *R*_0_ for both models, except with interventions applied to the the viral loads and therefore the dynamics. Prediction of R without direct simulation is described in Supplemental Text S1.

## Data Availability

Simulation code is available via GitHub.

https://github.com/LarremoreLab/covid_surveillance_testing

## Acknowledgements

The authors wish to thank the BioFrontiers Institute IT HPC group. This work was supported by grants NIH F32 AI145112 (James Burke), NIH F30 AG063468 (Evan Lester), MURIW911NF1810208 (Bryan Wilder, Milind Tambe), an NIH directors DP5 award 1DP50D028145-01 (Michael Mina), and the Howard Hughes Medial Institute (Roy Parker).

## Supplemental Figures and Tables

**Figure S1:**
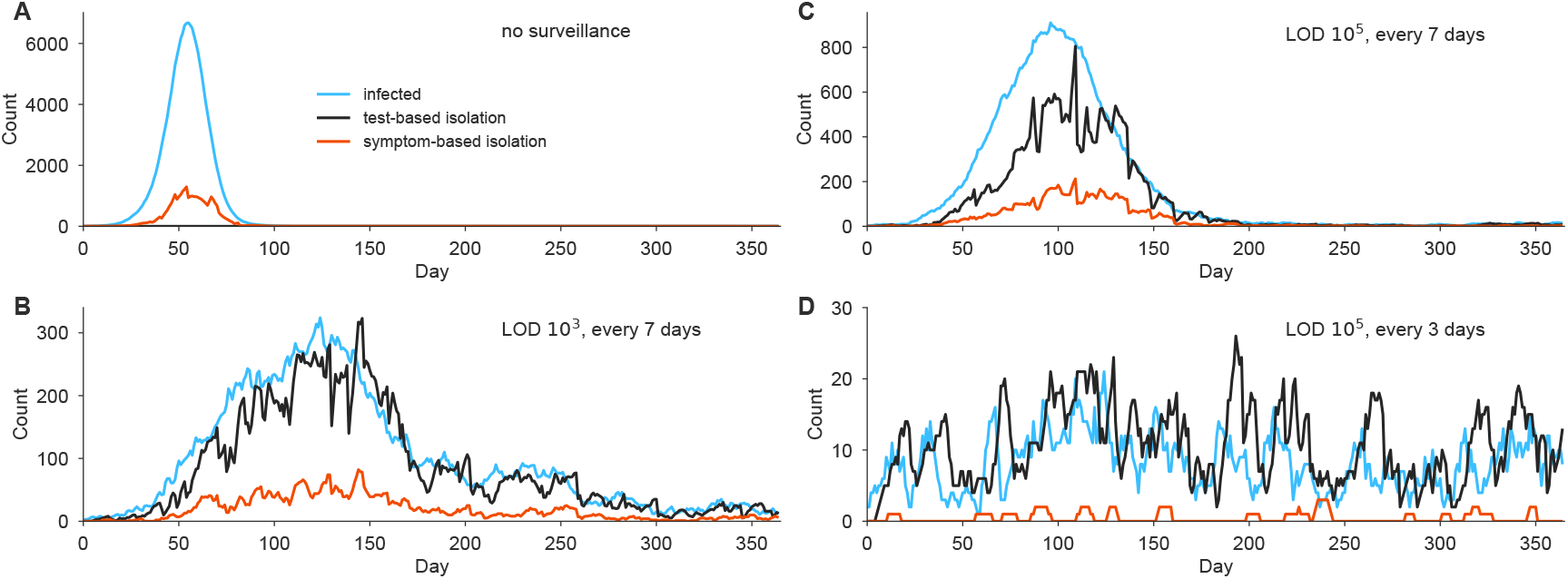
Sample simulation trajectories from fully mixed model. Simulation trajectories show the number of infected individuals in a population of N = 20,000 with a constant rate of external infection set to *1/N* per person per day, i.e. around 1 imported case per day. Infections (blue), test-based isolation (black), and symptom-based isolation (red) are shown for four scenarios, with *R*_0_ = 2.5. (A) No surveillance. (B) Weekly testing at LOD 10^3^. (C) Weekly testing at LOD 10^5^. (D) Testing every 3 days with LOD 10^5^. Note the variation in the vertical axis scales. The model is fully described in Methods.

**Figure S2:**
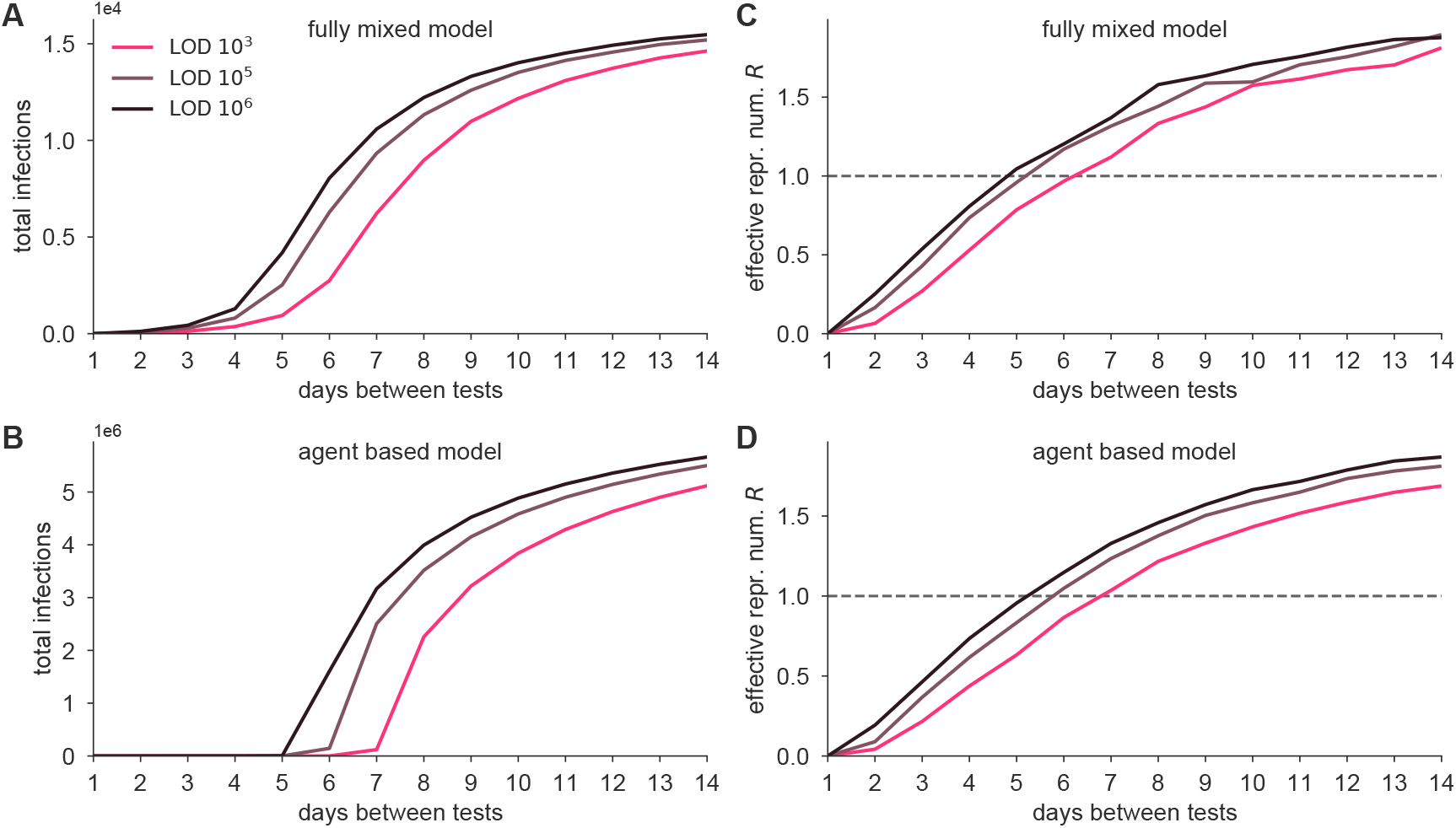
Epidemiological model outcomes for various test LODs and frequencies. The fully mixed model (top row) and agent based model (bottom row) were simulated (Methods) with various test frequencies, ranging from daily to once every 14 days, and with LODs of 10^3^, 10^5^, and 10^6^. Modeling results show mean outcomes from 50 independent simulations at each point, expressed as (A, B) total infections and (C, D) effective reproductive number *R*, from a baseline of *R*_0_ = 2.5. For the fully mixed model, only secondary infections are shown, excluding imported infections. Total population sizes were *N* = 2 x 10^4^ for the fully mixed model and 8.4 x 10^6^ for the agent based model. Dashed lines indicate *R* = 1 for reference.

**Figure S3:**
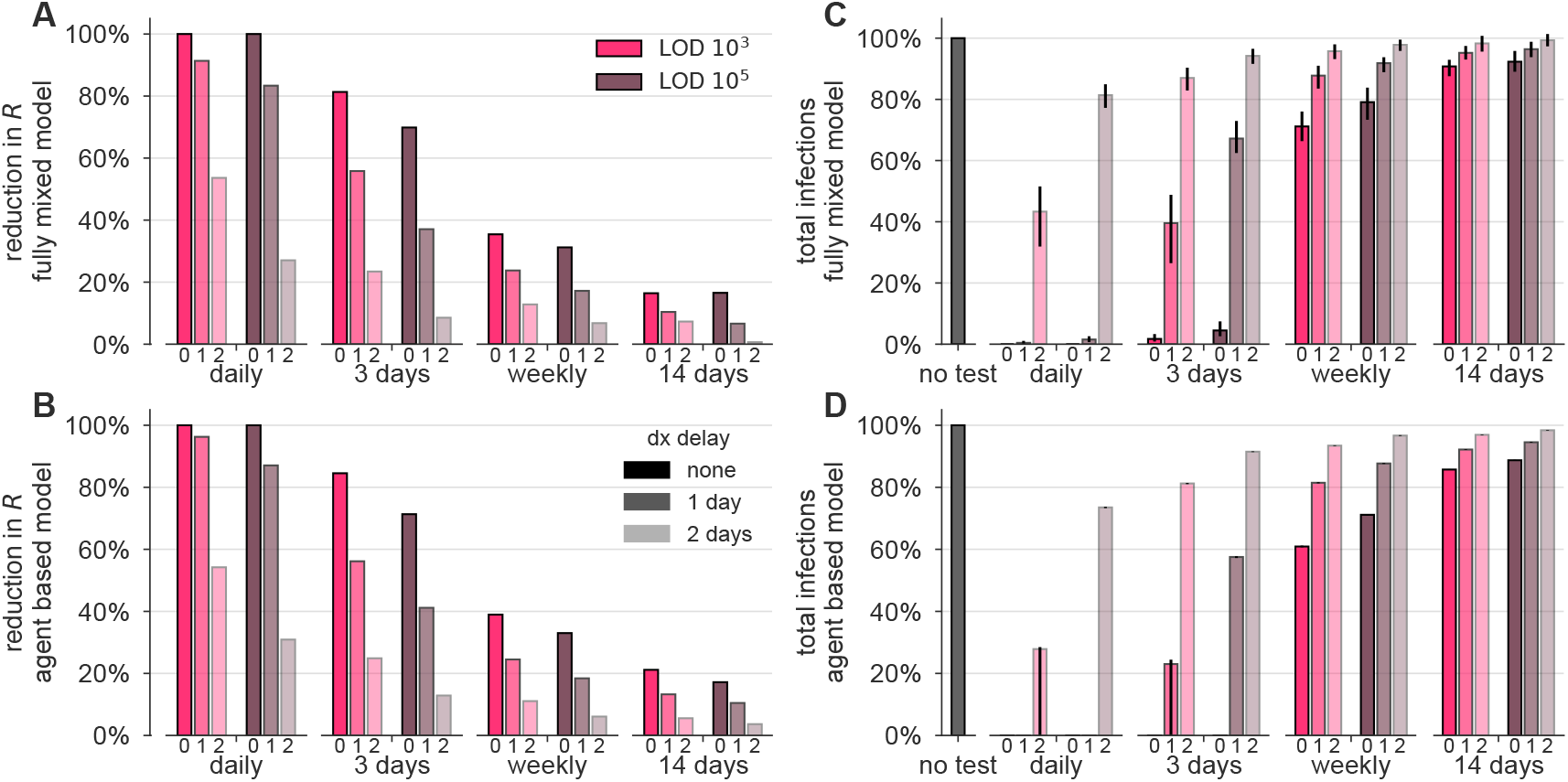
Delays in reporting decrease the epidemiological impact of surveillance-driven isolation (proportional infectiousness) This figure presents results from simulations which were identical to those shown in the main text Figure 4, but in which infectiousness was assumed to be *directly proportional* to viral load. Compare with threshold (binary) infectiousness in Fig. S4 and log-proportional infectiousness in Fig 4. See Methods.

**Figure S4:**
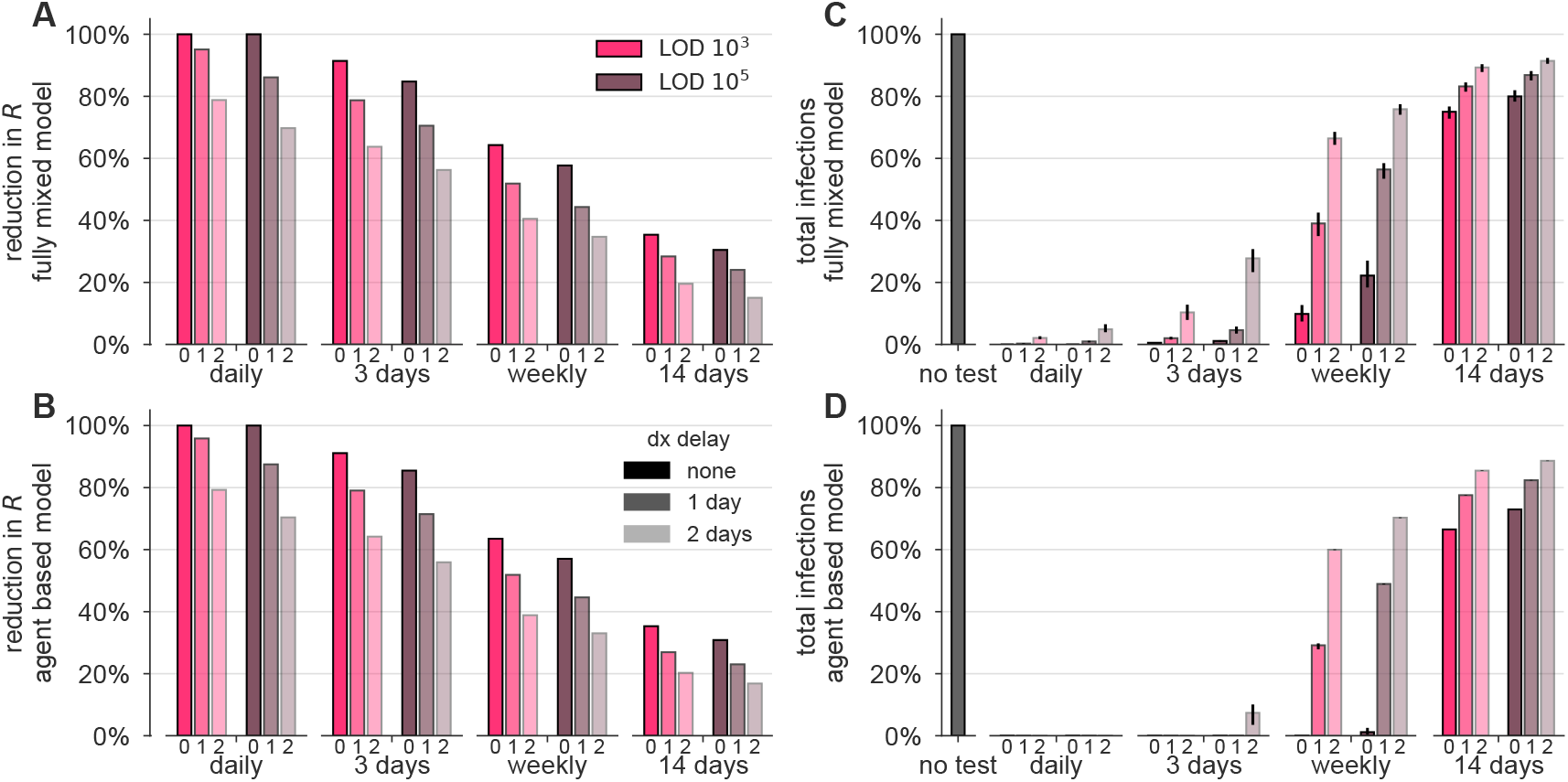
Delays in reporting decrease the epidemiological impact of surveillance-driven isolation (threshold infectiousness) This figure presents results from simulations which were identical to those shown in the main text Figure 4, but in which infectiousness was assumed to be *binary*, i.e. no infectiousness below 10^6^ and equal infectiousness for any viral load above 10^6^. Compare with proportional infectiousness in Fig. S3 and log-proportional infectiousness in Fig 4. See Methods.

**Figure S5:**
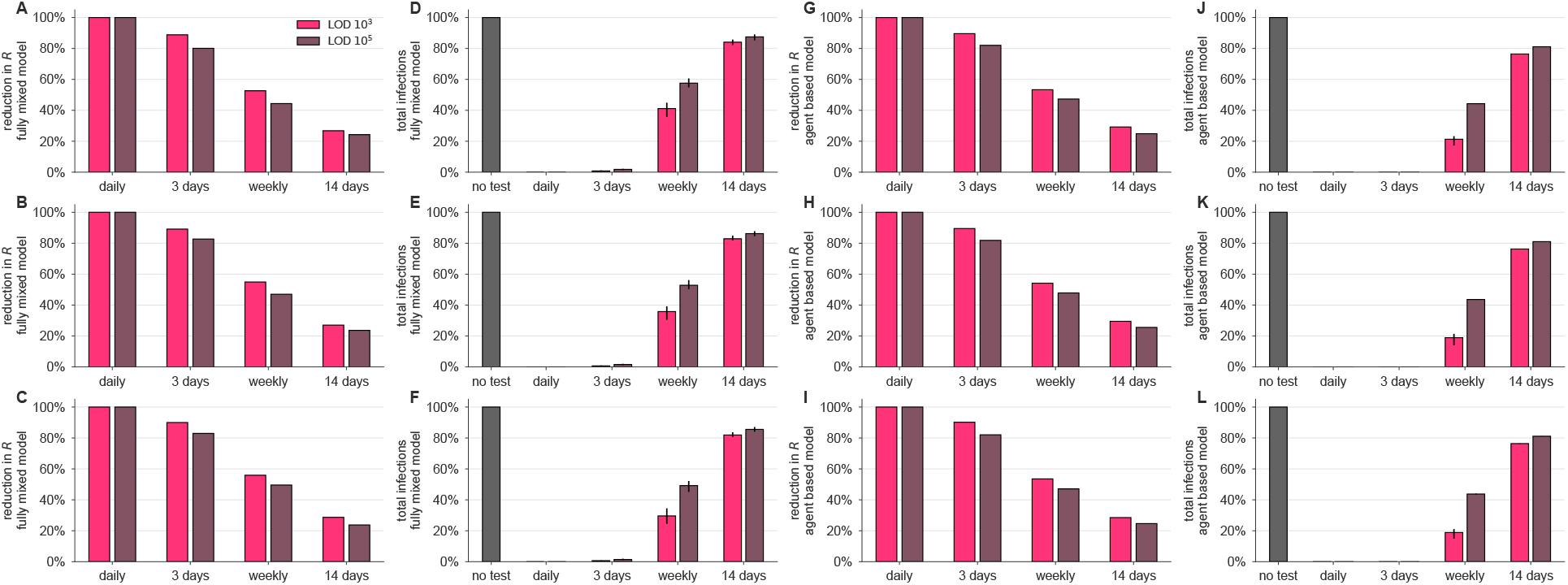
Robustness of surveillance effectiveness to the fraction of individuals who are symptomatic. (A-F) Results from fully-mixed simulations and (G-L) agent-based simulations using an asymptomatic rate of 50% (top row), 65% (middle row; identical to main text Fig 2), and 80% (bottom row).

**Figure S6:**
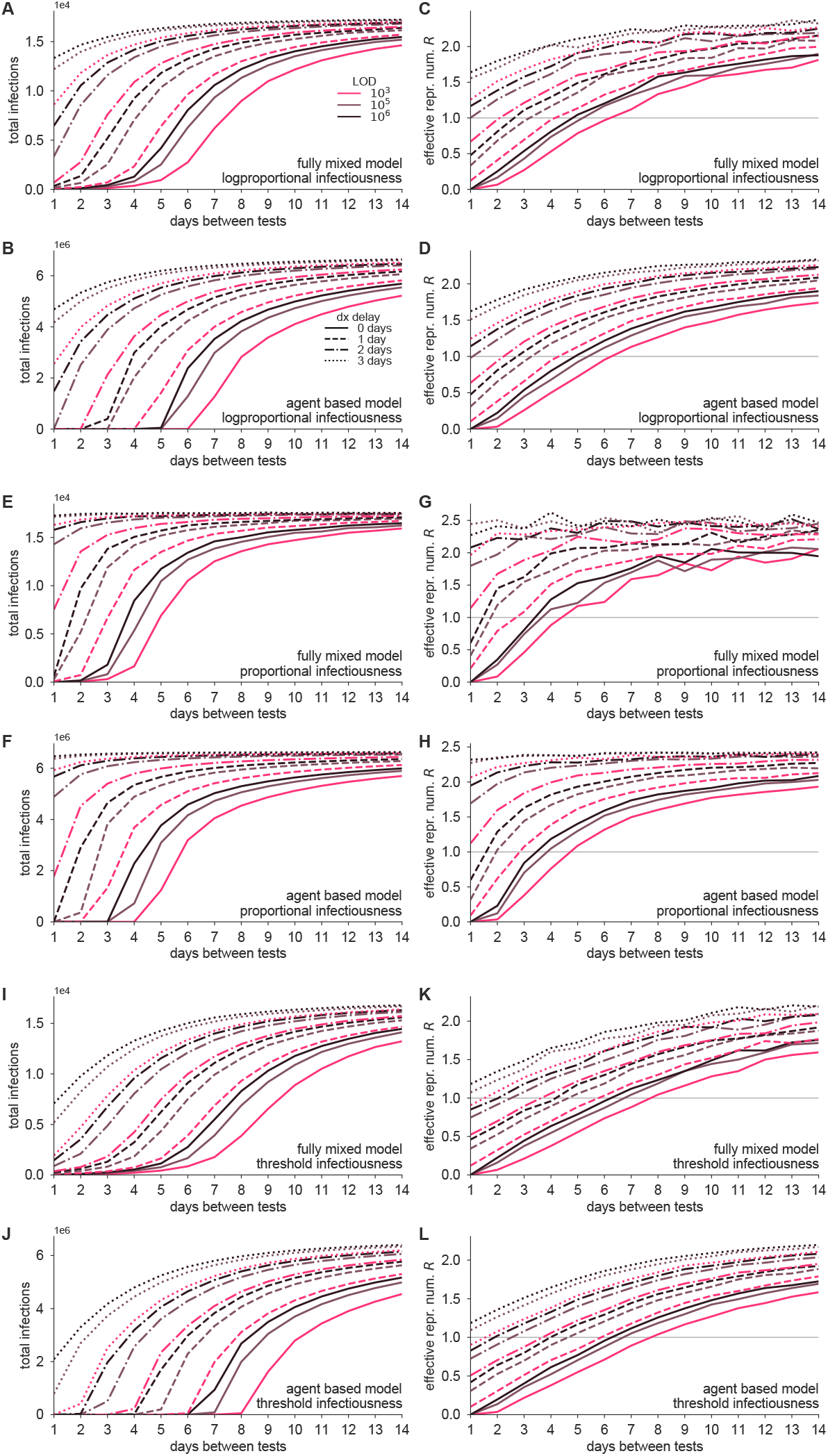
Epidemiological model outcomes for various test LODs, frequencies, infectiousness functions, and with reporting delays. The fully mixed model and agent based model were simulated (Methods) with various test frequencies, ranging from daily to once every 14 days, with LODs of 10^3^, 10^5^, and 10^6^, and with delays of 0, 1, 2, or 3 days, for log-proportional, proportional, and threshold infectiousness functions (see Methods). Legends in panels A and B indicate LODs and delays, and in-plot annotations describe various conditions. Modeling results show mean outcomes from 50 independent simulations at each point, expressed as total infections and effective reproductive number R, from a baseline of *R*_0_ = 2.5. For the fully mixed model, only secondary infections are shown, excluding imported infections. Total population sizes were N = 2 x 10^4^ for the fully mixed model and 8.4 x 10^6^ for the agent based model. A horizontal line indicates R =1 for reference.

**Figure S7:**
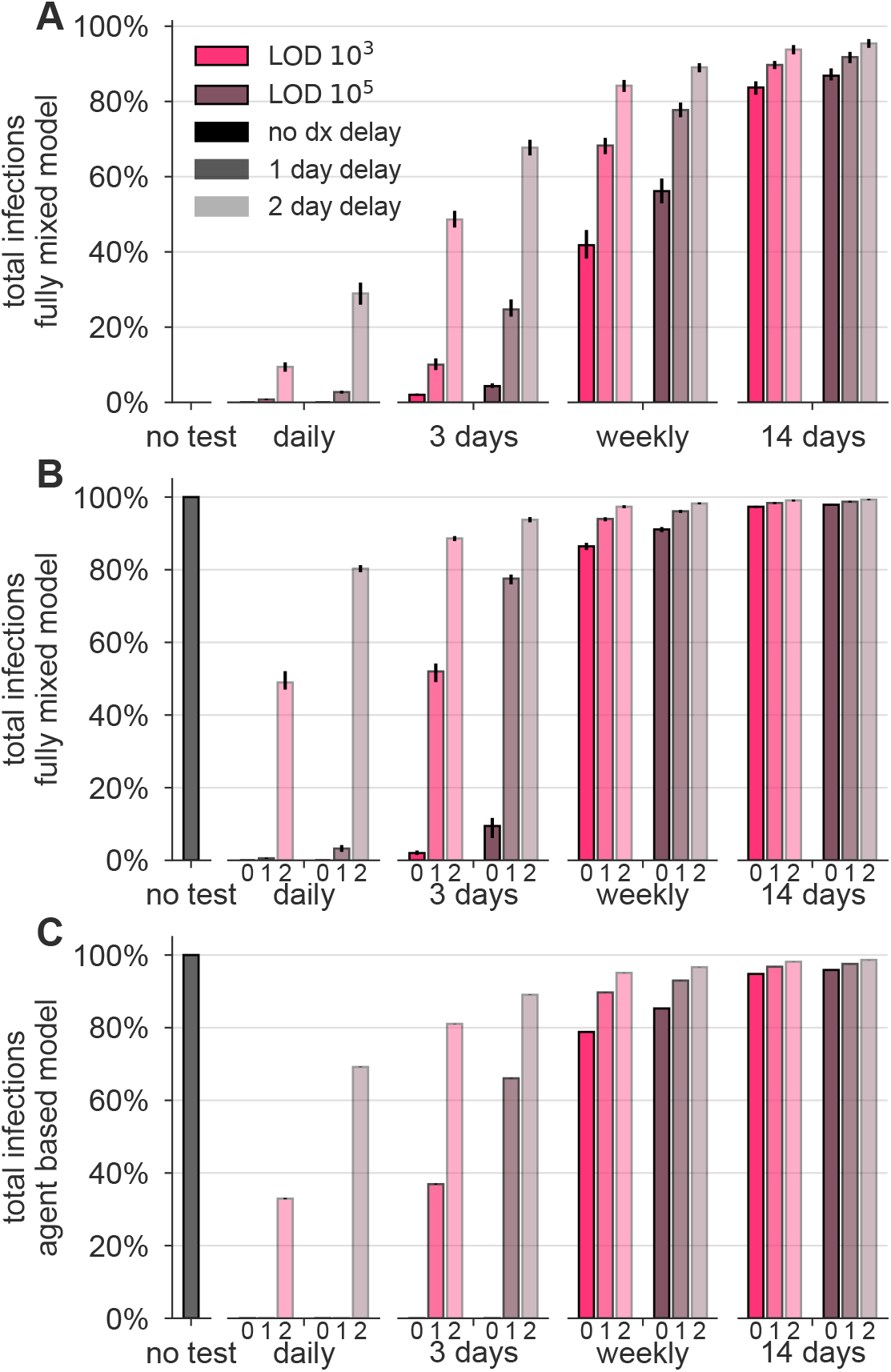
Robustness of surveillance effectiveness to epidemiological model parameters. (A) Results from the fully-mixed simulation with a tripled rate of external infection, i.e. 3/N per person per day. (B) Results from the fully mixed simulation with *R*_0_ doubled, i.e. *R*_0_ = 5. (C) Results from the agent-based simulation with *R*_0_ doubled, i.e.

**Figure S8:**
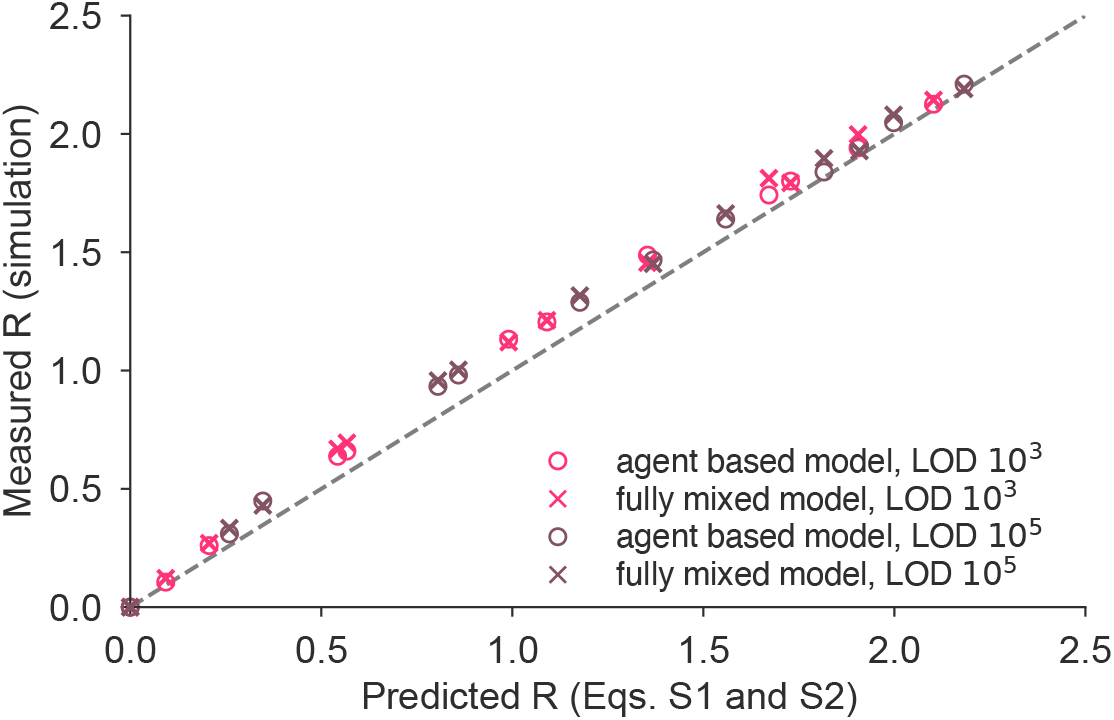
Predicted and simulated impact of surveillance on the reproductive number *R*. Mathematical predictions of the reproductive number *R* (see Equation (S1) in Supplemental Text S1) are scattered against their empirical measurements for the simulations shown in the main text (Figs. 2 and 4). Pearson’s r = 0.998, p < 10^-6^.

**Figure S9:**
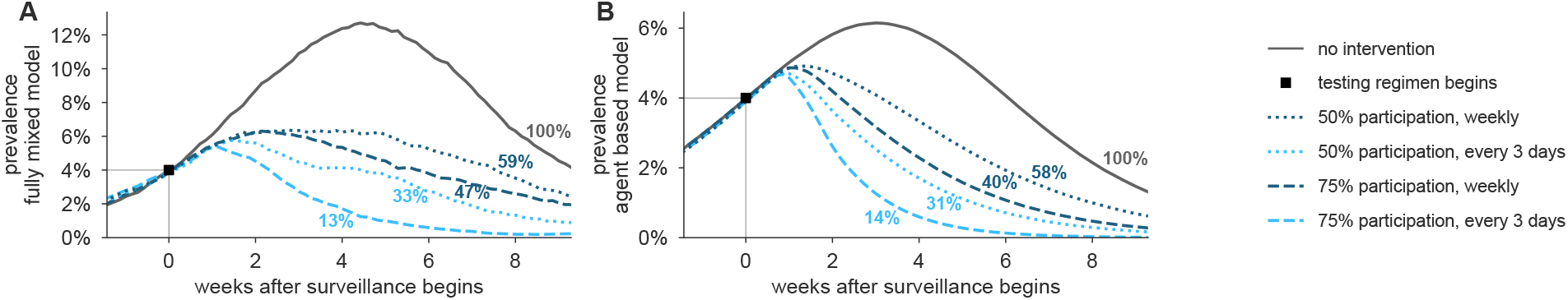
Surveillance testing suppresses an ongoing epidemic using a test with LOD 10^6^. Widespread testing and isolation of infected individuals drives prevalence downward for both (A) the fully-mixed compartmental model and (B) the agent based model. Time-series of prevalence, measured as the total number of infectious individuals, are shown for no intervention (solid) and surveillance testing scenarios (various dashed; see legend). Surveillance testing began only when prevalence reached 4% (box), and time series are shifted such that testing begins at t = 0. Scenarios show the impact of a test with LOD 10^6^, no delay in results, and with 10% of samples assumed to be incorrectly collected (and therefore negative) to reflect decreased sensitivity incurred at sample collection in a mass testing scenario. Annotations show total number of post-intervention infections, as a percentage of the no-intervention scenario, labeled as 100%. See Fig. 5 for identical simulations using a test with LOD 10^5^.

## Supplemental Text

### S1 Predicting the impact of surveillance testing on *R*

The impact of surveillance on the reproductive number can be estimated by considering the ratio of population infectiousness with surveillance testing to population infectiousness with no surveillance testing. However, note that the impact of a surveillance testing policy may depend on two additional factors.

First, not all individuals may wish to participate in a testing program. Let the fraction of individuals who participate be given by *ϕ*.

Second, a test may produce a false negative result *unrelated* to its limit of detection—for instance due to an improperly collected sample. Let se be the test sensitivity, in the particular sense of the probability of correctly diagnosing an individual as positive when that person’s viral load should, in principle, have provided a sufficiently high RNA concentration to be detectable.

Let *f*_0_ be the total infectiousness removed with no testing policy, i.e. due to symptom-driven self isolation. Let *f*_test_(se) be the fraction of total infectiousness removed with a chosen testing policy, inclusive of symptom-driven self isolation, as well as the test sensitivity se introduced above.

Both *f*_0_ and *f*_test_(se) can be estimated rapidly via Monte Carlo by drawing trajectories and applying a surveillance policy to them in which a fraction 1 – *se* positive tests are discarded uniformly at random. In the main text, we found that estimating these values using 10, 000 randomly drawn trajectories was sufficient to produce stable estimates.

Under the assumption of statistical independence between an individual’s participation or refusal, viral load, and *se*, we can approximate the reproductive number as

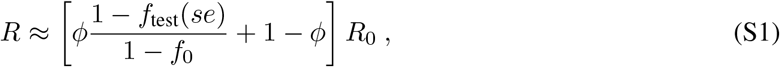

which simply expresses a weighted combination of removed infectiousness via surveillance testing participation and no test. Intuitively, note that if there is complete refusal to participate (*ϕ* = 0) or an entirely ineffective test (*f*_test_(*se*) = *f*_0_), then *R* ≈ *R*_0_, as expected.

## References

[1] Melissa M Arons, Kelly M Hatfield, Sujan C Reddy, Anne Kimball, Allison James, Jesica R Jacobs, Joanne Taylor, Kevin Spicer, Ana C Bardossy, Lisa P Oakley, et al. Presymptomatic SARS-CoV-2 infections and transmission in a skilled nursing facility. New England Journal of Medicine, 2020.

[2] Desmond Sutton, Karin Fuchs, Mary D’alton, and Dena Goffman. Universal screening for SARS-CoV-2 in women admitted for delivery. New England Journal of Medicine, 2020.

[3] Daniel P Oran and Eric J Topol. Prevalence of asymptomatic SARS-CoV-2 infection: A narrative review. Annals of Internal Medicine, 2020.

[4] Seyed M Moghadas, Meagan C Fitzpatrick, Pratha Sah, Abhishek Pandey, Affan Shoukat, Burton H Singer, and Alison P Galvani. The implications of silent transmission for the control of covid-19 outbreaks. Proceedings of the National Academy of Sciences, 117(30):17513-17515, 2020.

[5] Nicholas C Grassly, Margarita Pons-Salort, Edward P K Parker, Peter J White, Neil M Ferguson, and “The Imperial College COVID-19 Response Team”. Comparison of molecular testing strategies for covid-19 control: a mathematical modelling study. The Lancet Infectious Diseases, 2020.

[6] Chantal BF Vogels, Anderson F Brito, Anne Louise Wyllie, Joseph R Fauver, Isabel M Ott, Chaney C Kalinich, Mary E Petrone, Marie-Louise Landry, Ellen F Foxman, and Nathan D Grubaugh. Analytical sensitivity and efficiency comparisons of SARS-CoV-2 qRT-PCR assays. medRxiv, 2020.

[7] Daniel J Butler, Christopher Mozsary, Cem Meydan, David Danko, Jonathan Foox, Joel Rosiene, Alon Shaiber, Ebrahim Afshinnekoo, Matthew MacKay, Fritz J Sedlazeck, et al. Shotgun transcriptome and isothermal profiling of sars-cov-2 infection reveals unique host responses, viral diversification, and drug interactions. bioRxiv, 2020.

[8] Viet Loan Dao Thi, Konrad Herbst, Kathleen Boerner, Matthias Meurer, Lukas PM Kremer, Daniel Kirrmaier, Andrew Freistaedter, Dimitrios Papagiannidis, Carla Galmozzi, Megan L Stanifer, et al. A colorimetric rt-lamp assay and lamp-sequencing for detecting sars-cov-2 rna in clinical samples. Science Translational Medicine, 12(556), 2020.

[9] Nicholas R Meyerson, Qing Yang, Stephen Kyle Clark, Camille L Paige, Will T Fattor, Alison R Gilchrist, Arturo Barbachano-Guerrero, and Sara L Sawyer. A community-deployable sars-cov-2 screening test using raw saliva with 45 minutes sample-to-results turnaround. medRxiv, 2020.

[10] Coronavirus (COVID-19) update: FDA informs public about possible accuracy concerns with Abbott ID NOW Point-of-Care Test. https://www.fda.gov/news-events/press-announcements/coronavirus-covid-19-update-fda-informs-public-about-possible-accuracy-concerns-abbott-id-now-point, May 14, 2020.

[11] Kendra Quicke, Emily Gallichote, Nicole Sexton, Michael Young, Ashley Janich, Gregory Gahm, Elizabeth J Carlton, Nicole Ehrhart, and Gregory D Ebel. Longitudinal surveillance for SARS-CoV-2 RNA among asymptomatic staff in five colorado skilled nursing facilities: Epidemiologic, virologic and sequence analysis. medRxiv, 2020.

[12] Roman Wolfel, Victor M Corman, Wolfgang Guggemos, Michael Seilmaier, Sabine Zange, Marcel A Muller, Daniela Niemeyer, Terry C Jones, Patrick Vollmar, Camilla Rothe, et al. Virological assessment of hospitalized patients with COVID-2019. Nature, 581(7809):465-469, 2020.

[13] Bernard La Scola, Marion Le Bideau, Julien Andreani, Van Thuan Hoang, Clio Grimaldier, Philippe Colson, Philippe Gautret, and Didier Raoult. Viral RNA load as determined by cell culture as a management tool for discharge of SARS-CoV-2 patients from infectious disease wards. European Journal of Clinical Microbiology & Infectious Diseases, 39(6):1059, 2020.

[14] Soren Alexandersen, Anthony Chamings, and Tarka Raj Bhatta. SARS-CoV-2 genomic and subgenomic RNAs in diagnostic samples are not an indicator of active replication. medRxiv, 2020.

[15] Charles GB Caraguel, Henrik Stryhn, Nellie Gagne, Ian R Dohoo, and K Larry Hammell. Selection of a cutoff value for real-time polymerase chain reaction results to fit a diagnostic purpose: analytical and epidemiologic approaches. Journal of Veterinary Diagnostic Investigation, 23(1):2-15, 2011.

[16] Adrian Ruiz-Villalba, Elizabeth van Pelt-Verkuil, Quinn D Gunst, Jan M Ruijter, and Maurice JB van den Hoff. Amplification of nonspecific products in quantitative polymerase chain reactions (qPCR). Biomolecular detection and quantification, 14:7-18,2017.

[17] PJ Klasse. Molecular determinants of the ratio of inert to infectious virus particles. In Progress in molecular biology and translational science, volume 129, pages 285-326. Elsevier, 2015.

[18] Muge Cevik, Matthew Tate, Oliver Lloyd, Alberto Enrico Maraolo, Jenna Schafers, and Antonia Ho. Sars-cov-2, sars-cov-1 and mers-cov viral load dynamics, duration of viral shedding and infectiousness: a living systematic review and meta-analysis. medRxiv, 2020.

[19] Amber M Smith and Alan S Perelson. Influenza A virus infection kinetics: quantitative data and models. Wiley Interdisciplinary Reviews: Systems Biology and Medicine, 3(4):429-445, 2011.

[20] Mathilde Richard, Adinda Kok, Dennis de Meulder, Theo M Bestebroer, Mart M Lamers, Nisreen MA Okba, Martje Fentener van Vlissingen, Barry Rockx, Bart L Haagmans, Marion PG Koopmans, et al. SARS-CoV-2 is transmitted via contact and via the air between ferrets. bioRxiv, 2020.

[21] Xi He, Eric HY Lau, Peng Wu, Xilong Deng, Jian Wang, Xinxin Hao, Yiu Chung Lau, Jessica Y Wong, Yujuan Guan, Xinghua Tan, et al. Temporal dynamics in viral shedding and transmissibility of COVID-19. Nature Medicine, 26(5):672-675, 2020.

[22] Zhuang Shen, Fang Ning, Weigong Zhou, Xiong He, Changying Lin, Daniel P Chin, Zonghan Zhu, and Anne Schuchat. Superspreading sars events, beijing, 2003. Emerging Infectious Diseases, 10(2):256, 2004.

[23] Joseph Sriyal Malik Peiris, Chung-Ming Chu, Vincent Chi-Chung Cheng, KS Chan, IFN Hung, Leo LM Poon, Kin-Ip Law, BSF Tang, TYW Hon, CS Chan, et al. Clinical progression and viral load in a community outbreak of coronavirus-associated sars pneumonia: a prospective study. The Lancet, 361(9371):1767-1772, 2003.

[24] Zheng Zhang, Tongyang Xiao, Yanrong Wang, Jing Yuan, Haocheng Ye, Lanlan Wei, Haiyan Wang, Xuejiao Liao, Shen Qian, Zhaoqin Wang, et al. Early viral clearance and antibody kinetics of COVID-19 among asymptomatic carriers. medRxiv, 2020.

[25] Enrico Lavezzo, Elisa Franchin, Constanze Ciavarella, Gina Cuomo-Dannenburg, Luisa Barzon, Claudia Del Vecchio, Lucia Rossi, Riccardo Manganelli, Arianna Loregian, Nicolo Navarin, etal, Suppression of a sars-cov-2 outbreak in the italian municipality of vo’. Nature, pages 1-5, 2020.

[26] Nguyen Van Vinh Chau, Vo Thanh Lam, Nguyen Thanh Dung, Lam Minh Yen, Ngo Ngoc Quang Minh, Nghiem My Ngoc, Nguyen Tri Dung, Dinh Nguyen Huy Man, Lam Anh Nguyet, Nguyen Thi Han Ny, et al. The natural history and transmission potential of asymptomatic sars-cov-2 infection. medRxiv, 2020.

[27] Xudan Chen, Yang Zhang, Baoyi Zhu, Jianwen Zeng, Wenxin Hong, Xi He, Jingfeng Chen, Haipeng Zheng, Shuang Qiu, Ying Deng, et al. Associations of clinical characteristics and antiviral drugs with viral rna clearance in patients with covid-19 in guangzhou, china: a retrospective cohort study. medRxiv, 2020.

[28] Zhiliang Hu, Ci Song, Chuanjun Xu, Guangfu Jin, Yaling Chen, Xin Xu, Hongxia Ma, Wei Chen, Yuan Lin, Yishan Zheng, et al. Clinical characteristics of 24 asymptomatic infections with covid-19 screened among close contacts in nanjing, china, Science China Life Sciences, 63(5):706-711, 2020.

[29] Rongrong Yang, Xien Gui, and Yong Xiong. Comparison of clinical characteristics of patients with asymptomatic vs symptomatic coronavirus disease 2019 in wuhan, china, JAMA Network Open, 3(5):e2010182-e2010182, 2020.

[30] Bryan Wilder, Marie Charpignon, Jackson A Killian, Han-Ching Ou, Aditya Mate, Shahin Jabbari, Andrew Perrault, Angel Desai, Milind Tambe, and Maimuna S Majumder. Modeling between-population variation in COVID-19 dynamics in hubei, lombardy, and new york city. Available at SSRN 3564800, 2020.

[31] Corey M Peak, Rebecca Kahn, Yonatan H Grad, Lauren M Childs, Ruoran Li, Marc Lipsitch, and Caroline O Buckee. Individual quarantine versus active monitoring of contacts for the mitigation of COVID-19: a modelling study. The Lancet Infectious Diseases, 2020.

[32] Adam J Kucharski, Petra Klepac, Andrew Conlan, Stephen M Kissler, Maria Tang, Hannah Fry, Julia Gog, John Edmunds, CMMID COVID-19 Working Group, et al. Effectiveness of isolation, testing, contact tracing and physical distancing on reducing transmission of SARS-CoV-2 in different settings. *medRxiv*, 2020.

[33] Yicheng Fang, Huangqi Zhang, Jicheng Xie, Minjie Lin, Lingjun Ying, Peipei Pang, and Wenbin Ji. Sensitivity of chest ct for COVID-19: comparison to RT-PCR. Radiology, page 200432, 2020.

[34] Anne Louise Wyllie, John Fournier, Arnau Casanovas-Massana, Melissa Campbell, Maria Tokuyama, Pavithra Vijayakumar, Bertie Geng, M Catherine Muenker, Adam J Moore, Chantal BF Vogels, et al. Saliva is more sensitive for SARS-CoV-2 detection in COVID-19 patients than nasopharyngeal swabs. Medrxiv, 2020.

[35] Elizabeth T Chin, Benjamin Q Huynh, Matthew Murrill, Sanjay Basu, and Nathan C Lo. Frequency of routine testing for covid-19 in high-risk environments to reduce workplace outbreaks. medRxiv, 2020.

[36] A David Paltiel, Amy Zheng, and Rochelle P Walensky. Assessment of sars-cov-2 screening strategies to permit the safe reopening of college campuses in the united states. JAMA network open, 3(7):e2016818-e2016818, 2020.

[37] David Holtz, Michael Zhao, Seth G. Benzell, Cathy Y. Cao, Mohammad Amin Rahimian, Jeremy Yang, Jennifer Allen, Avinash Collis, Alex Moehring, Tara Sowrirajan, Dipayan Ghosh, Yunhao Zhang, Paramveer S. Dhillon, Christos Nicolaides, Dean Eckles, and Sinan Aral. Interdependence and the cost of uncoordinated responses to covid-19. Proceedings of the National Academy of Sciences, 117(33):19837-19843, 2020.

[38] Tracy A Lieu, G Thomas Ray, Nicola P Klein, Cindy Chung, and Martin Kulldorff. Geographic clusters in underimmunization and vaccine refusal. Pediatrics, 135(2):280-289, 2015.

[39] Stephen Kissler, Nishant Kishore, Malavika Prabhu, Dena Goffman, Yaakov Beilin, Ruth Landau, Cynthia Gyamfi-Bannerman, Brian Bateman, Daniel Katz, Jonathan Gal, Angela Bianco, Joanne Stone, Daniel Larremore, Caroline Buckee, and Yonatan Grad. Reductions in commuting mobility correlate with geographic differences in sars-cov-2 prevalence in new york city. Nature Communications (in press), 2020.

[40] Stephen A Lauer, Kyra H Grantz, Qifang Bi, Forrest K Jones, Qulu Zheng, Hannah R Meredith, Andrew S Azman, Nicholas G Reich, and Justin Lessler. The incubation period of coronavirus disease 2019 (COVID-19) from publicly reported confirmed cases: estimation and application. Annals of internal medicine, 172(9):577-582, 2020.

[41] Esteban Abelardo Hernandez Vargas and Jorge X Velasco-Hernandez. In-host modelling of covid-19 kinetics in humans. medRxiv, 2020.

[42] James M Burke, Clovis R Bass, Rodney P Kincaid, Emin T Ulug, and Christopher S Sullivan. The murine polyomavirus microrna locus is required to promote viruria during the acute phase of infection. Journal of virology, 92(16):e02131-17, 2018.

[43] Abishek Chandrashekar, Jinyan Liu, Amanda J Martinot, Katherine McMahan, Noe B Mercado, Lauren Peter, Lisa H Tostanoski, Jingyou Yu, Zoltan Maliga, Michael Nekorchuk, et al. SARS-CoV-2 infection protects against rechallenge in rhesus macaques. Science, 2020.

[44] Jingyou Yu, Lisa H Tostanoski, Lauren Peter, Noe B Mercado, Katherine McMahan, Shant H Mahrokhian, Joseph P Nkolola, Jinyan Liu, Zhenfeng Li, Abishek Chandrashekar, et al. Dna vaccine protection against SARS-CoV-2 in rhesus macaques. Science, 2020.

[45] Benny Borremans, Amandine Gamble, KC Prager, Sarah K Helman, Abby M McClain, Caitlin Cox, Van Savage, and James O Lloyd-Smith. Quantifying antibody kinetics and RNA shedding during early-phase SARS-CoV-2 infection. medRxiv, 2020.

[46] Jeroen J.A. van Kampen, David A.M.C. van de Vijver, Pieter L.A. Fraaij, Bart L. Haagmans, Mart M. Lamers, Nisreen Okba, Johannes P.C. van den Akker, Henrik Endeman, Diederik A.M.P.J. Gommers, Jan J. Cornelissen, Rogier A.S. Hoek, Menno M. van der Eerden, Dennis A. Hesselink, Herold J. Metselaar, Annelies Verbon, Jurriaan E.M. de Steenwinkel, Georgina I. Aron, Eric C.M. van Gorp, Sander van Boheemen, Jolanda C. Voermans, Charles A.B. Boucher, Richard Molenkamp, Marion P.G. Koopmans, Corine Geurtsvankessel, and Annemiek A. van der Eijk. Shedding of infectious virus in hospitalized patients with coronavirus disease-2019 (covid-19): duration and key determinants. medRxiv, 2020.

[47] Hitoshi Kawasuji, Yusuke Takegoshi, Makito Kaneda, Akitoshi Ueno, Yuki Miyajima, Koyomi Kawago, Yasutaka Fukui, Yoshihiko Yoshida, Miyuki Kimura, Hiroshi Yamada, et al. Viral load dynamics in transmissible symptomatic patients with COVID-19. medRxiv, 2020.

[48] Kelvin Kai-Wang To, Owen Tak-Yin Tsang, Wai-Shing Leung, Anthony Raymond Tam, Tak-Chiu Wu, David Christopher Lung, Cyril Chik-Yan Yip, Jian-Piao Cai, Jacky Man-Chun Chan, ThomasShiu-Hong Chik, et al. Temporal profiles of viral load in posterior oropharyngeal saliva samples and serum antibody responses during infection by SARS-CoV-2: an observational cohort study. The Lancet Infectious Diseases, 2020.

[49] Jin Yong Kim, Jae-Hoon Ko, Yeonjae Kim, Yae-Jean Kim, Jeong-Min Kim, Yoon-Seok Chung, Heui Man Kim, Myung-Guk Han, So Yeon Kim, and Bum Sik Chin. Viral load kinetics of SARS-CoV-2 infection in first two patients in korea. Journal of Korean medical science, 35(7), 2019.

[50] Ai Tang Xiao, Yi Xin Tong, and Sheng Zhang. Profile of RT-PCR for SARS-CoV-2: a preliminary study from 56 COVID-19 patients. Clinical Infectious Diseases, 2020.

[51] Yang Liu, Li-Meng Yan, Lagen Wan, Tian-Xin Xiang, Aiping Le, Jia-Ming Liu, Malik Peiris, Leo LM Poon, and Wei Zhang. Viral dynamics in mild and severe cases of COVID-19. The Lancet Infectious Diseases, 2020.

[52] Minnesota Population Center. Integrated public use microdata series, international: Version 7.2 [dataset], 2019. https://doi.org/10.18128/D020.V7.2.

[53] Yang Liu, Rosalind Eggo, and Adam Kucharski. Secondary attack rate and superspreading events for SARS-CoV-2. The Lancet, 2020.

[54] Kiesha Prem, Alex Cook, and Mark Jit. Projecting social contact matrices in 152 countries using contact surveys and demographic data. PLoS Computational Biology, 13(9):e1005697, 2017.

